# Biomarkers of LRRK2 and lysosomal dysfunction in Progressive Supranuclear Palsy

**DOI:** 10.1101/2025.11.24.25340830

**Authors:** Louise-Kristine Nielsen, Joshua LI Frost, David P Vaughan, Raquel Real, Riona Fumi, Marte Theilmann Jensen, Megan Hodgson, Eleanor J. Stafford, Lesley Wu, Olaf Ansorge, Annelies Quaegebeur, Kieren SJ Allinson, Thomas T Warner, Zane Jaunmuktane, Anjum Misbahuddin, P Nigel Leigh, Boyd CP Ghosh, Kailash P Bhatia, Alistair Church, Christopher Kobylecki, Michele TM Hu, James B Rowe, Alan A Shomo, Danielle L Graham, Omar S Mabrouk, Huw R Morris, Esther M Sammler, Edwin Jabbari

## Abstract

**Background:** Common and rare genetic variants in LRRK2 have been linked with sporadic and familial Parkinson’s disease (PD). Recently, we discovered that common genetic variation near the LRRK2 locus determined survival in progressive supranuclear palsy (PSP). Our study aimed to explore biomarkers of LRRK2 and lysosomal dysfunction in PSP.

**Methods:** Immunoblotting was used to measure total LRRK2 and LRRK2-dependent Rab10 phosphorylation at Threonine73 (pRab10^Thr73^) in neutrophil and monocyte samples from PSP and control participants. Urine samples were applied to a multiplexed assay to quantitate bis(monoacylglycerol)phosphate (BMP) species as markers of lysosomal dysfunction. CSF samples from a wider cohort of PSP and control participants were applied to a stable isotope standards and capture by anti-peptide antibodies assay to measure total LRRK2 and pRab10^Thr73^ levels. LRRK2 genotypes (rs76904798 and rs2242367) and 1-year change in PSP rating scale scores were obtained.

**Results:** 61 PSP and 34 control participants were included. Total urine 22:6-BMP levels were higher in PSP vs. control samples (p=0.04) and correlated with CSF total LRRK2 levels (r=0.49, p=0.04). In PSP, carriers of the alternate allele (CT and TT genotypes) at the LRRK2 PD risk variant, rs76904798, had higher levels of CSF total LRRK2 vs. CC genotype (p=0.02). A similar but non-significant trend was observed for the LRRK2 PSP survival variant, rs2242367 (p=0.08). Baseline monocyte total LRRK2 levels predicted 1-year change in the PSP rating scale score (p=0.008).

**Conclusions:** Biochemically defined lysosomal dysfunction is evident in PSP. Genetic and biochemical stratification may identify PSP patients that would benefit from LRRK2-targetting therapies.

## Introduction

Progressive supranuclear palsy (PSP) and Parkinson’s disease (PD) are neurodegenerative parkinsonian disorders with no effective disease-modifying therapies. Although they have overlapping clinical features, the two disorders are pathologically distinct with PD being characterised by the presence of alpha-synuclein related neuronal Lewy body pathology and PSP being characterised by neuronal and astrocytic 4-repeat tau pathology (**1**).

Around 1% of all PD and 5% of familial PD cases are due to rare pathogenic mutations in the leucine-rich repeat kinase 2 (LRRK2) gene (LRRK2-PD), with the G2019S variant being the most common cause of familial PD in populations of European ancestry (**2**). Additionally, common genetic variation at LRRK2 (rs76904798) is a well-established risk locus for sporadic PD (**3**). LRRK2 mutations have rarely been associated with PSP, with pathogenic LRRK2 variants identified in 0.3% of a large post-mortem PSP cohort (**4**). However, we have recently shown that common variation near the LRRK2 locus (rs2242367) is a genetic determinant of survival in a genome-wide association study of over 1000 pathologically diagnosed PSP participants (**5**).

Interestingly, the neuropathology of LRRK2-PD is unclear, as typical Lewy body pathology is absent in around half of all cases that come to post-mortem, with previous studies highlighting the common presence of tau pathology (**6**). This is also reflected in alpha-synuclein seed amplification (a-syn SAA) studies of PD where around 30-40% of LRRK2-PD participants are a-syn SAA negative (**7**).

LRRK2 encodes a multi-domain protein with catalytic GTPase and kinase domains. In PD, pathogenic LRRK2 variants cluster in these two catalytic domains and increase LRRK2 kinase activity with subsequent hyperphosphorylation of its endogenous targets, a subgroup of RabGTPases. The resulting activation leads to pathogenic downstream effects, including impaired vesicle trafficking, lysosomal dysfunction and neuroinflammation related to microglial activation, all of which may contribute to PD pathogenesis (**8**). As such, LRRK2 kinase inhibitors and LRRK2 lowering therapies, including anti-sense oligonucleotides (ASO), are currently being trialled in both sporadic PD and LRRK2-PD (**9**).

Translational assays have been developed to probe LRRK2 function in peripheral blood neutrophils and monocytes where LRRK2 is highly expressed. Quantitative immunoblotting and more sensitive mass-spectrometry assays have enabled the quantification of total and phosphorylated levels of both LRRK2 and a subgroup of LRRK2 phosphorylated RabGTPases, including Rab10 at the threonine 73 residue (pRab10^Thr73^), in neutrophil and monocyte samples as markers of LRRK2 kinase activity (**10**). Previous studies have shown significantly higher neutrophil and monocyte levels of Rab10 phosphorylation in LRRK2-PD (R1441G mutation) vs. controls, a non-significant trend towards similar results in LRRK2-PD (G2019S mutation) vs. controls, and no difference in levels between sporadic PD and controls (**11**). Similarly, urine levels of bis(Monoacylglycerol)Phosphate (BMP) species, a marker of lysosomal dysfunction, have been shown to be raised in both manifesting and non-manifesting LRRK2-PD vs. sporadic PD whereas there is no significant difference in levels between sporadic PD and controls (**12**). Furthermore, in the Parkinson’s Progression Markers Initiative (PPMI) cohort, urine BMP levels were shown to elevated in PD participants who were carriers of the GBA1 N409S variant vs. controls, but this was much smaller than the three-to-seven-fold elevation seen in LRRK2-PD vs. controls (**13**).

In this study, we have explored whether blood and CSF biomarkers of the LRRK2 pathway and urine biomarkers of lysosomal dysfunction are: 1) deranged in PSP vs. controls; 2) determined by LRRK2 single nucleotide polymorphisms, i.e. protein quantitative trait loci (pQTL); 3) predict clinical disease progression in PSP.

## Methods

### Patient consent and cohorts

All participants gave written informed consent for the use of their medical records and biosamples for research purposes, including the analysis of DNA. Participants from the PROSPECT-UK study (Queen Square Research Ethics Committee 14/LO/1575) who fulfilled at least “possible” clinical diagnostic criteria for PSP were included in this study (**14**)(**15**). Age and sex-matched neurologically normal controls from the PROSPECT-UK study were also included. Post-mortem evaluation was conducted on a subset of participants with a clinical diagnosis of PSP.

### Clinical data collection

The following clinical variables and measures were collected from PSP participants: sex; age at symptom onset; age and disease duration at baseline assessment; baseline and 1-year follow-up scores for the PSP rating scale (PSPRS), Movement Disorder Society-Unified Parkinson’s Disease Rating Scale part III (MDS-UPDRS-III) in the off state, and Montreal Cognitive Assessment (MoCA).

### Neutrophil and monocyte LRRK2 and pRab10 detection by quantitative immunoblotting

For each participant, neutrophil and monocyte samples were isolated from fresh whole blood samples, divided into two and treated with either DMSO (inactive) or MLi-2 (LRRK2 inhibitor), and stored at -70°C within 3 hours of blood sampling as previously described (**10**). Multiplexed immunoblotting was carried out using stored neutrophil and monocyte samples as per established protocols **(Supplementary Methods)**. The quantified signal intensities of the total protein bands were normalised to those of the housekeeping protein (total LRRK2/GAPDH and total Rab10/GAPDH), whereas the phosphorylated species were normalised to the total protein signal intensities (pSer935/total LRRK2 and pThr73/total Rab10). All measurements were then normalised against an inter-gel control, which was run on all gels to control for variability.

### CSF LRRK2 and pRab10 detection by SISCAPA

A subset of the PSP and control participants that had undergone fresh blood and urine sampling also had a lumbar puncture to obtain CSF for LRRK2 and pRab10 testing. Importantly, these CSF assays are able to be performed on samples stored at -70°C with no pre-processing required and so CSF samples previously obtained from additional PSP and control PROSPECT-UK participants were also included. However, we only used CSF samples that had not previously undergone freeze-thaw cycles.

CSF (500µl) was spiked with 1X RIPA (Millipore Sigma 20-188). Samples were then spiked with 1pg of LRRK2 stable isotope labelled peptide K_13C_(_6_)_15N_(_2_) AEEGDLLVNPDQPR_13C_(_6_)_15N_(_2_) and 100fg of pRab10 Thr73 stable isotope labelled peptide FHTITTSYYR_13C_(_6_)_15N_(_2_) as internal standards. Samples were then digested for 1.5 hours at 37°C with 10µg of TPCK-treated trypsin (Millipore Sigma T1426).

N241A/34 LRRK2 antibody (Antibodies Inc. 75-253) and MJF-R20 pRab8A antibody (Abcam ab230260) were biotinylated then immobilized on AssayMAP 5µL streptavidin SA-W cartridges (Agilent Technologies G5496-60010) at a ratio of 1µg of each antibody per cartridge. Digested samples were then loaded onto the AssayMAP Bravo for peptide immunocapture. Eluted samples were loaded onto an Evosep One LC system running the 40SPD Whisper method coupled to a Thermo EASY-Spray PepMap RSLC C18 column (Thermo Scientific ES904). Mass spectrometry was performed on a Thermo Exploris 480 operating in PRM mode. Targets included *m/z* 560.9566 [M+3H]^3+^ (native LRRK2 peptide), *m/z* 566.9641 [M+3H]^3+^ (LRRK2 internal standard), *m/z* 456.8710 [M+3H]^3+^ (native pRab10 Thr73 phosphopeptide) and *m/z* 460.2071 [M+3H]^3+^ (pRab10 Thr73 internal standard). PRM scans were collected at a resolution of 120,000 with AGC target set to standard (1×10^6^) and maximum injection time set to automatic detection. HCD collision energy was optimized at 20% and 25% for the LRRK2 and pRab10 peptide pairs respectively. Quantitation and data analysis were performed in SkyLine 24.1.0.199.

### Urine BMP detection

For each participant, fresh urine samples were obtained and centrifuged for 15 minutes at 2500 x g and 4°C before 1ml of supernatant was stored at -70°C within 30 minutes of urine sampling. Stored urine samples were analysed at Nextcea Inc. (Woburn, MA) using a multiplexed UPLC-MS/MS method to simultaneously quantitate BMP species (total di-22:6-BMP, 2,2’ di-22:6-BMP, 2,3’ di-22:6-BMP, 3,3’ di-22:6-BMP, total di-18:1-BMP, 2,2’ di-18:1-BMP, 2,3’ di-18:1-BMP, 3,3’ di-18:1-BMP) as per established protocols (**12**). Measured concentrations of urine BMPs (ng/mL) were divided by the concentration of urine creatinine and reported as ng/mg creatinine.

### Genetic testing

DNA samples from PSP and control participants underwent PCR-based (KASP) genotyping at LGC Biosearch Technologies for the LRRK2-G2019S variant (rs34637584), the LRRK2 PD risk variant (rs76904798), and the LRRK2 PSP survival variant (rs2242367). Whole Genome Sequencing (WGS) and Illumina Neurobooster Array (NBA) (**16**) genotyping was performed on a subset of DNA samples that had initially undergone KASP genotyping at LGC Biosearch Technologies. WGS data was available via release 10 of the Global Parkinson’s Genetics Program (https://gp2.org). We screened for variants defined as pathogenic, likely pathogenic or risk factor according to ClinVar (https://www.ncbi.nlm.nih.gov/clinvar/) across genes known to be associated with PD/Parkinsonism **(Supplementary Table 1)**. NBA data was screened for pathogenic variants across the same list of genes known to be associated with PD/Parkinsonism as in the WGS data analysis **(Supplementary Table 1)**.

### Statistical analyses

All statistical analyses were carried out using R version 4.5.1.

Urine BMP and neutrophil, monocyte and CSF LRRK2 and Rab10 data were screened for extreme outliers, which were defined as values above the third quartile plus 3 times the interquartile range (IQR) or values below the first quartile minus 3 times the IQR. All results were individually checked for exclusion against published ranges for each biomarker.

Group level analyses of neutrophil and monocyte biomarkers were done using multinomial logistic regression via the nnet package. Logistic regression analyses were also used to model: 1) group level differences between urine and CSF biomarker levels; 2) neutrophil, monocyte and CSF biomarker levels stratified by LRRK2 PD risk (rs76904798, C:C vs. T:T/T:C) and PSP survival (rs2242367, G:G vs. A:A/A:G) SNP status within each disease group separately. All of the above comparisons were visualised using violin plots from the ggplot2 package, and accompanying odds ratios and 95% confidence intervals were also calculated. All of the above models adjusted for sex, age and disease duration at testing except for disease vs. control group models which only adjusted for sex and age at testing.

Assessment of correlation between neutrophil, monocyte, urine and CSF biomarkers in PSP and control groups was done using Spearman correlation with pairwise complete observations, visualised on heat maps. Statistically significant correlations were modelled linearly and adjusted for covariates.

In the PSP group, linear regression was used to model whether baseline levels of biomarkers predicted 1-year change in the PSP rating scale score. Significant relationships were visualised as scatter plots with linear fit lines.

Statistical significance was defined as p-value < 0.05 for all analyses aside from the exploratory progression analysis where the threshold for p-value significance was set using the Benjamini-Hochberg correction method (**17**) for multiple testing with a false discovery rate of 5%.

## Data availability

The raw data used for analyses in this study will be considered for sharing in anonymised format by request of a qualified investigator to the corresponding author for purposes of replicating procedures and results.

## Results

61 PSP participants and 34 age and sex-matched controls from the PROSPECT-UK study were included in this study **(Table 1)**. LGC genotyping was available for 50/61 (82%) PSP and 26/34 (76%) control participants. WGS data was available for 23/61 (38%) PSP and 5/34 (15%) control participants. No pathogenic variants were identified in PSP or control participants via WGS data. NBA data was available for an additional 18 PSP and 14 control participants that had not undergone WGS. This did not reveal any pathogenic variants.

**Table 1:** Clinical profile of the LRRK2 biomarker cohort. Corticobasal degeneration: CBD, Progressive Supranuclear Palsy: PSP, PSP-Richardson syndrome: PSP-RS, PSP-parkinsonism: PSP-P, PSP-progressive gait freezing: PSP-PGF, PSP/Corticobasal syndrome overlap: PSP/CBS, PSP-speech and language disorder: PSP-SL, PSP-frontal: PSP-F, number: n, standard deviation: SD, PSP rating scale: PSPRS, Movement Disorder Society-Unified Parkinson’s Disease Rating Scale part III: MDS-UPDRS III, Montreal Cognitive Assessment: MoCA. Fisher’s exact test used for group comparisons of sex distributions. Group comparisons of continuous variables were done using linear regression that was either unadjusted (*) or adjusted (^) for sex, age and disease duration at baseline (group comparisons of MoCA scores involving controls were only adjusted for sex and age at baseline). 12 = p<0.05 vs. control group.

|  | **Progressive supranuclear palsy (n=61)** | **Controls (n=34)** |
| --- | --- | --- |
| **Sex (% male)** | 56% | 38% |
| **Clinical subtype at baseline assessment (n)** | PSP-RS, n=37<br>PSP-P, n=12<br>PSP-PGF, n=4<br>PSP/CBS, n=3<br>PSP-F, n=3<br>PSP-SL, n=2 |  |
| **Mean age at symptom onset, years (SD) \*** | 64.8 (7.1) years |  |
| **Mean age at baseline assessment,** | 68.7 (6.9) years<br>□ | 61.8 (12.5) years |
| **years (SD) \*** |  |  |
| **Mean disease duration at baseline assessment, years (SD) \*** | 3.9 (2.0) years |  |
| **Mean PSPRS at baseline assessment, score (SD) ^** | 32.7 (12.4) |  |
| **Mean MDS-UPDRS III at baseline assessment, score (SD) ^** | 34.3 (13.6) |  |
| **Mean MoCA at baseline assessment, score (SD) ^** | 21.8 (4.5)<br>□ | 27.6 (1.6) |
| **Participants deceased at censoring (n), mean disease duration from symptom onset to death, years (SD)** | n=32<br>6.9 (2.8) years |  |
| **Primary pathological diagnosis in participants undergoing post-mortem (n)** | PSP, n=5<br>CBD, n=1 |  |

Fresh blood samples from 30 PSP and 30 control participants underwent neutrophil and monocyte extraction followed by treatment with DMSO and MLi-2 before being stored at -70°C. Quantitative immunoblotting was subsequently performed on stored neutrophil and monocyte samples. Two PSP monocyte samples were excluded from analyses as it was not possible to accurately quantify their immunoblotting bands which led to extreme outlier results. For the remaining neutrophil and monocyte samples, MLi-2 treated samples showed a depletion in the levels of pLRRK2^Ser935^ pRab10^Thr73^ which confirmed that the levels of LRRK2 and Rab10 phosphorylation in DMSO treated samples were mediated by LRRK2 kinase activity. We found no PSP vs. control group level differences in the levels of total and phosphorylated LRRK2 and Rab10 in DMSO treated neutrophil and monocyte samples **(Supplementary Figure 1)**. From the same set of participants, 29/30 PSP and 28/30 control participants provided urine samples which underwent processing before being stored at -70°C. One PSP patient was unable to provide a urine sample due to advanced disease and urinary incontinence, and urine samples from two controls were excluded due to improper processing prior to storage. BMP assay testing was performed on stored urine samples. One control sample was excluded from analyses due to the presence of extreme outliers in the BMP data. Group level analyses revealed significantly higher levels of total di-22:6-BMP as well as 3,3’ di-18:1-BMP isoform in PSP vs. controls (p<0.05) **(Figure 1) (Supplementary Table 2)**.

**Figure 1:**
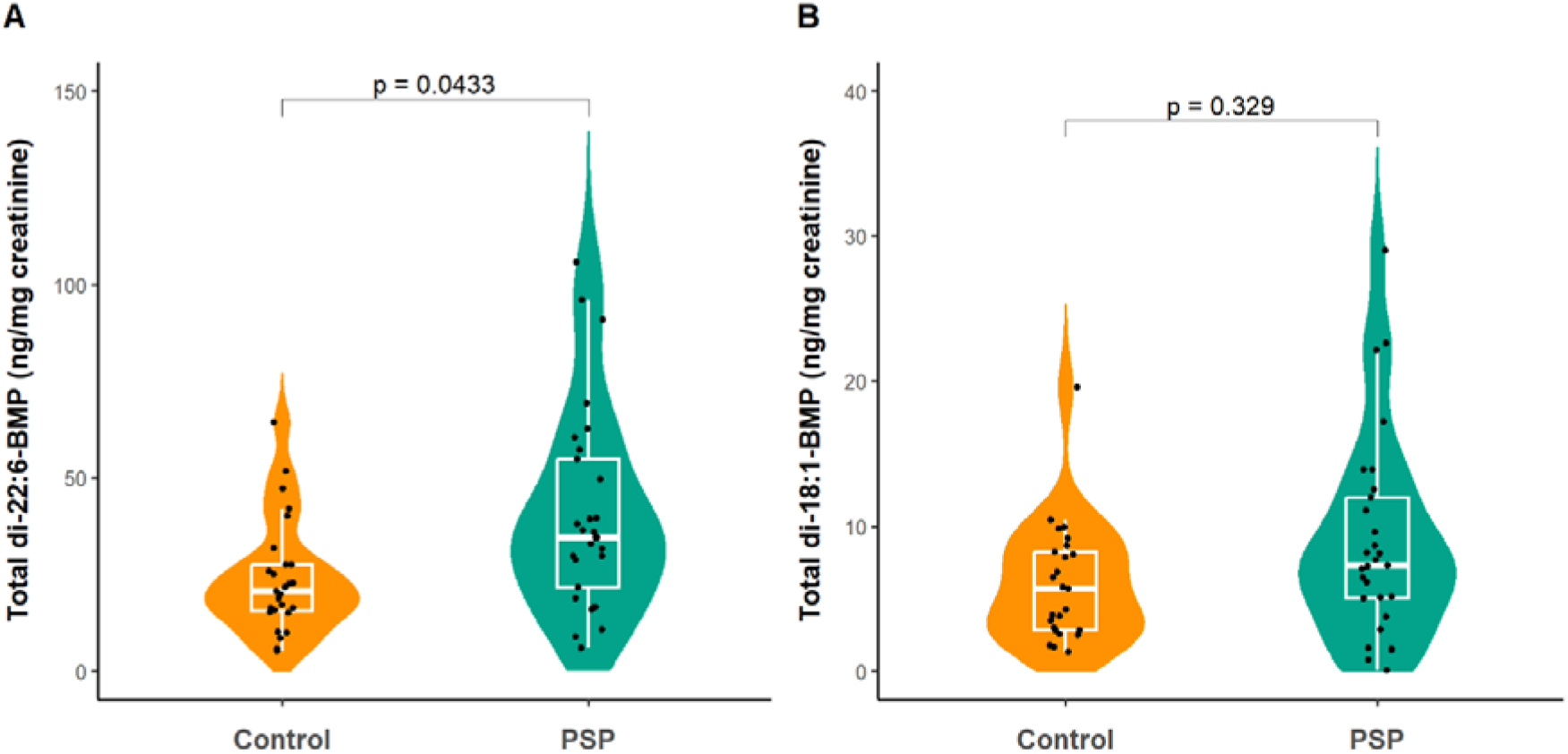
Urine total di-22:6 and total di-18:1-BMP levels in PSP and control participants. Group comparisons done using logistic regression that adjusted for sex and age at testing.

Stored CSF samples from 51 PSP and 9 control participants underwent testing for total LRRK2 and pRab10^Thr73^ levels. Group level analyses revealed no differences in CSF total LRRK2 levels **(Figure 2a-b)**. Using LGC genotype data, we then stratified our blood (neutrophil and monocyte) and CSF measures of total LRRK2 and pRab10 in each group separately by LRRK2 PD risk (rs76904798) and PSP survival (rs2242367) SNP status to look for state-specific pQTLs. This analysis revealed that in PSP, carriers of the alternate ‘T’ allele (CT and TT genotypes) at rs76904798 had higher levels of CSF total LRRK2 vs. CC genotype (p=0.02) **(Figure 2c)**. A non-significant (p=0.08) trend towards rs2242367 genotype status influencing CSF total LRRK2 levels was observed in the PSP group **(Figure 2d)**. There were no pQTL associations observed in: a) neutrophil and monocyte levels of total LRRK2 and pRab10; b) CSF levels of pRab10 **(Supplementary Table 3)**. Of note, in the 24 PSP and 8 control participants who had undergone both LGC genotyping and WGS, there was 100% concordance in the genotype results for rs76904798 and rs2242367.

**Figure 2:**
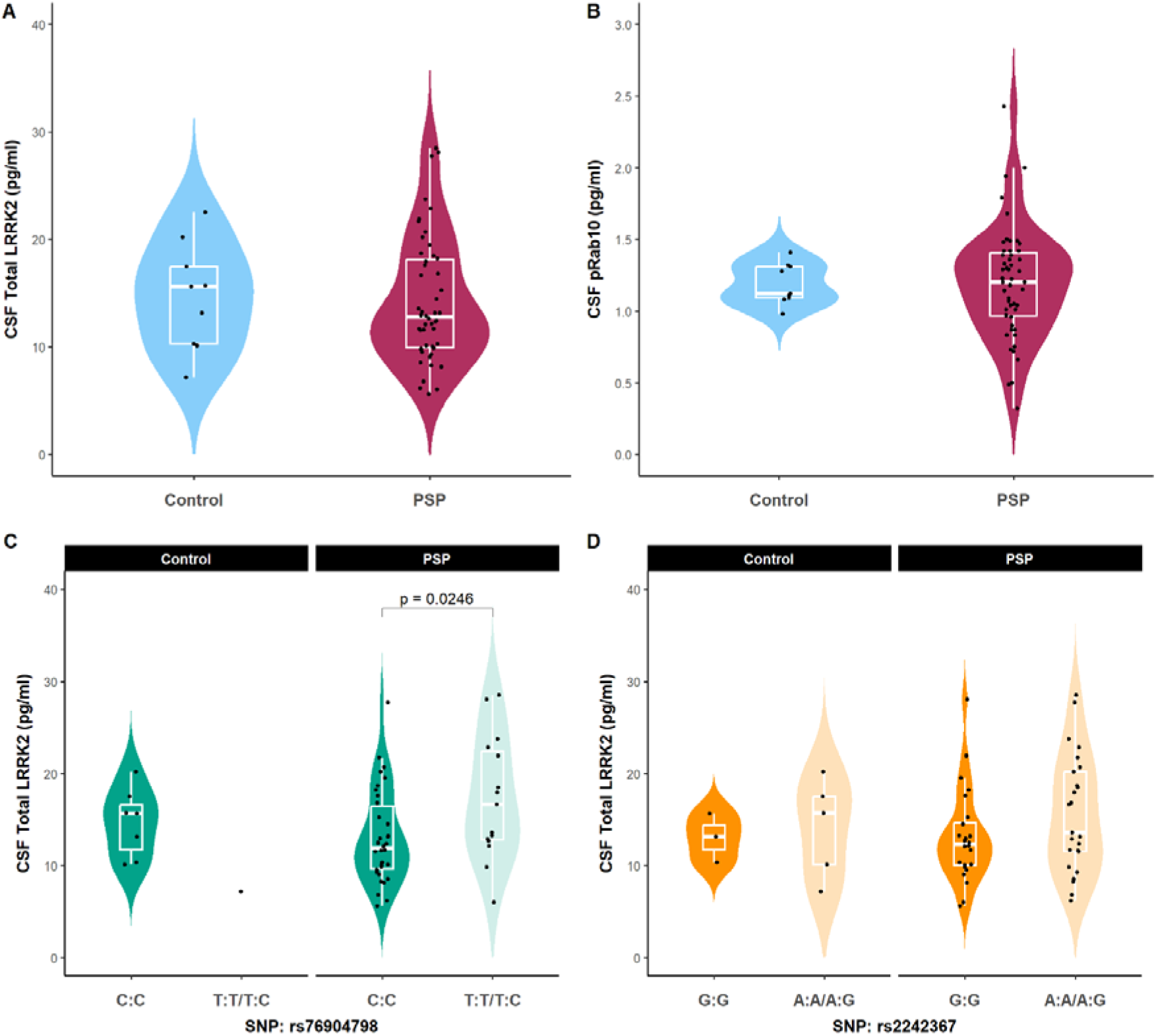
a-b) CSF total LRRK2 and pRab10 levels in PSP and control groups. Group comparisons were done using logistic regression that adjusted for sex, age and disease duration at testing (control group comparisons adjusted for sex and age at testing); c-d) CSF total LRRK2 levels in PSP and control groups stratified by rs76904798 and rs2242367 genotype status. Genotype group comparisons were done using logistic regression that adjusted for sex, age and disease duration at testing (control group comparisons adjusted for sex and age at testing) with GG (rs2242367) and CC (rs76904798) as reference groups.

We assessed for correlation between each of the blood, urine and CSF biomarkers in PSP and control groups respectively. Of note, this included 19 PSP and 3 control participants who had undergone testing for each of the blood, urine and CSF measures. In the PSP group, the only correlations that were statistically significant in linear regression analyses that adjusted for sex, age and disease duration at testing were urine total di-22:6 BMP vs. CSF LRRK2 (r=0.49, p=0.04) and CSF LRRK2 vs. CSF pRab10 (r=0.67, p<0.0001) **(Figure 3)**.

**Figure 3:**
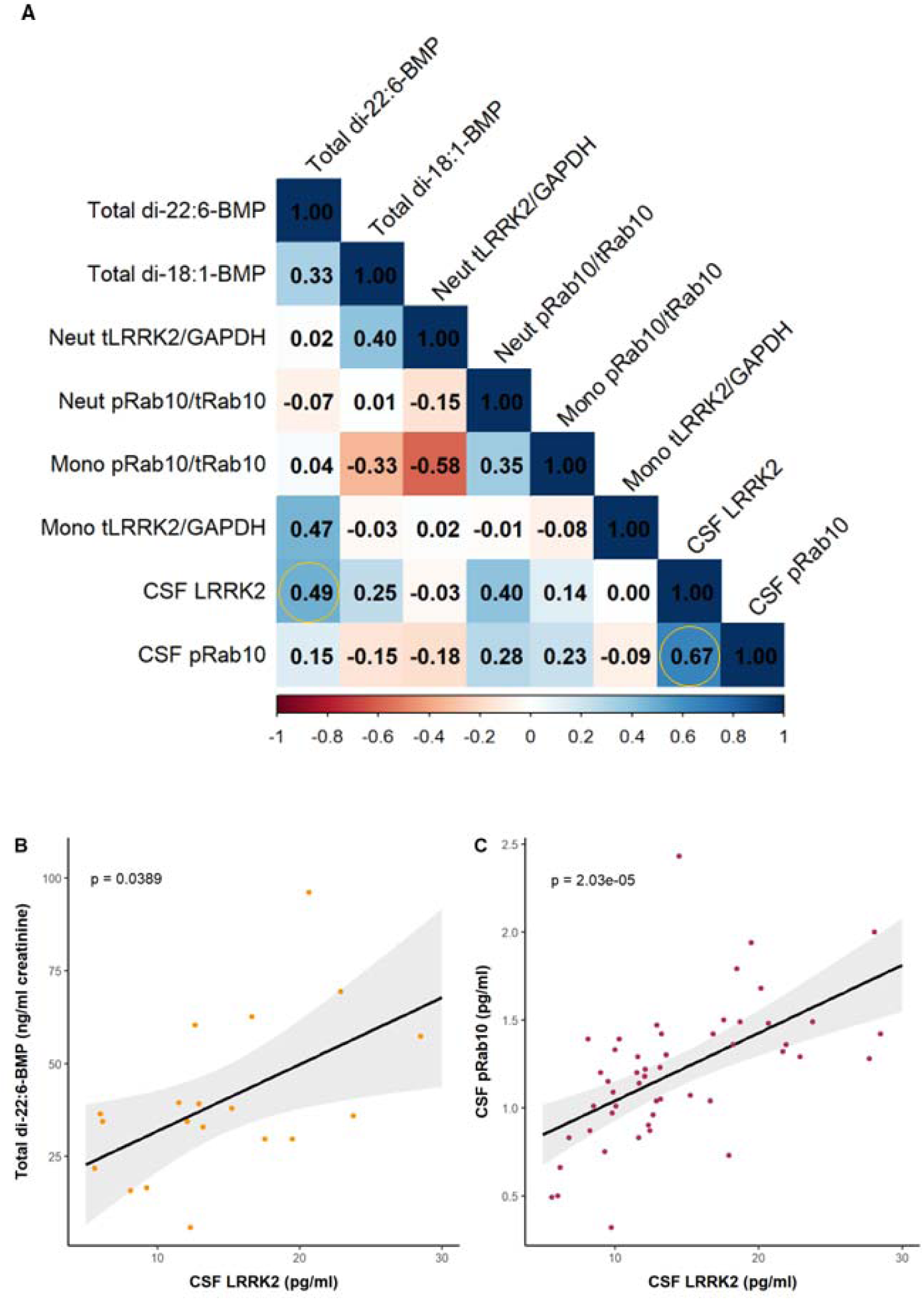
a) Heatmap of Spearman’s rho biomarker correlations in the PSP group with r values highlighted. Circles represent correlations that were also significant in the linear regression analyses; b-c) urine total di-22:6 BMP vs. CSF LRRK2 and CSF LRRK2 vs. CSF pRab10 linear plot. Linear regression analyses that adjusted for sex, age and disease duration at testing were used to generate p-values.

Of note, there were no significant correlations between neutrophil or monocyte LRRK2 vs. pRab10, neutrophil or monocyte LRRK2 vs. CSF LRRK2, or neutrophil or monocyte pRab10 vs. CSF pRab10. In the control group, the only correlation that was statistically significant in linear regression analyses that adjusted for sex and age at testing was neutrophil pRab10 vs. monocyte pRab10 **(Supplementary Figure 2)**.

Finally, in the PSP group we assessed whether baseline levels of the blood, CSF and urine measures predicted 1-year change in the PSP rating scale score via linear regression models that adjusted for sex, age and disease duration at baseline. Of note, in the PSP group, 1-year clinical rating scale scores were available in 12/30 (40%) of participants that had undergone baseline blood and urine testing and 28/51 (55%) of participants that had undergone baseline CSF testing. The average 1-year change in the PSP rating scale score was 12.0 points, standard deviation 10.3. The progression analyses revealed that baseline monocyte total LRRK2 levels predicted 1-year change in the PSP rating scale score (p=0.008) **(Figure 4) (Supplementary Table 4)**.

**Figure 4:**
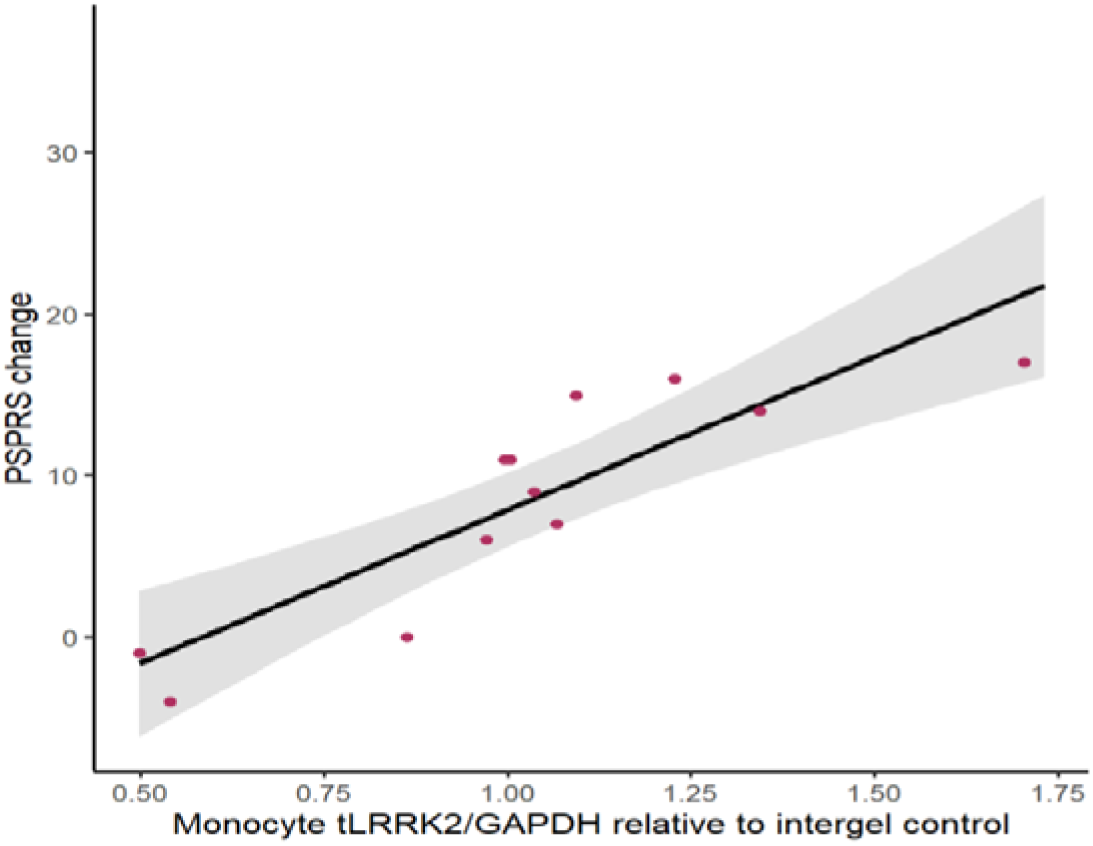
Baseline monocyte total LRRK2 vs.1 year change in the PSP rating scale score.

## Discussion

In this study we have comprehensively characterised the profile of fluid biomarkers of LRRK2 and lysosomal dysfunction in PSP and assessed their association with genetic variation at the LRRK2 locus and clinical disease progression.

First, we used an established quantitative immunoblotting assay and found no evidence of differences in neutrophil and monocytes levels of total and phosphorylated LRRK2 and Rab10 in PSP vs. controls. Future replication studies should prioritise more sensitive mass-spectrometry methods to quantify a range of LRRK2-dependent Rab proteins. Of note, a recent analysis of PSP brain tissue lysates and immunohistochemistry revealed that pRab12 labelled lysosomal-like granulovacuolar bodies in neurons and showed co-pathology with tau inclusions (**18**).

Although we did not observe any group-level differences in CSF total LRRK2 and pRab10 vs. controls, stratification of results by LRRK2 PD risk SNP (rs76904798) status revealed significantly higher levels of CSF total LRRK2 in risk allele (T) carriers vs. CC genotype. This suggests that a subset of PSP patients have genetically determined pathogenically raised LRRK2 levels and this occurs independently of pRab10 such that these patients may benefit from LRRK2 lowering therapies, including ASOs, as opposed to kinase inhibitors. This conclusion is further reinforced by our finding of baseline monocyte total LRRK2 levels predicting 1 year change in PSP rating scale scores in the PSP group. The effect of monocyte LRRK2 levels on disease progression may relate to peripheral immune response-driven neuroinflammation or be due to a specific effect in monocyte or microglial lineage cells (**19**). Furthermore, the therapeutic potential of targeting LRRK2 in PSP has been shown in a recent whole-genome CRISPR screen which highlighted the role of LRRK2 in controlling the endocytosis of monomeric tau in human neurons (**20**).

Of note, we have previously shown that common variation near the LRRK2 locus (rs2242367) determines survival in PSP, and carrying the alternate allele at rs2242367 was associated with reduced survival and increased expression of LRRK2 in whole blood (**5**). However, in this study we only observed a non-significant trend towards rs2242367 genotype status determining CSF total LRRK2 levels in PSP. It should be noted that whilst rs76904798 and rs2242367 are in low *r*² linkage disequilibrium, they are within 200kb of each other and have a high DD suggesting that they may be part of a haplotype block which determines disease progression in PSP. It is also possible that rs76904798 and rs2242367 have different effects on regulating LRRK2 expression in different cells and/or in different cellular states which may be relevant to the development of PD and PSP. In line with this, a recent study showed that rs76904798 status determines the level of microglial LRRK2 RNA expression in post-mortem brain tissue and in a patient induced pluripotent stem cell-derived microglia model (**21**).

We found significantly higher levels of urine BMP species in PSP vs. controls. Although raised urine BMP has previously been detected in LRRK2-PD, it has also been found in lysosomal storage disorders including Niemann-Pick type C disease such that it is considered a non-specific marker of lysosomal dysfunction (**22**).

However, in PSP, raised urine BMP levels may potentially be driven by LRRK2 dysfunction due to the positive correlation we observed between urine BMP and CSF LRRK2 levels, although further work is required to explore this hypothesis. In the PPMI cohort, urine BMP levels remained stable over two years in LRRK2-PD participants. Furthermore, in LRRK2-PD, GBA-PD and sporadic PD, baseline urine BMP levels did not predict disease progression as measured by striatal DaT imaging, MDS-UPDRS-III and MoCA. Altogether, these results suggested that urine BMP may be used as a target modulation biomarker in clinical trials for monogenic and sporadic PD, but not as a prognostic or disease progression biomarker (**13**).

The main limitation of this study was the small sample size and limited number of PSP participants that had 1-year clinical follow up data available which is why our progression analyses should be considered exploratory. Independent replication in larger genetically characterised PSP cohorts alongside sporadic PD and LRRK2-PD positive control groups will be essential to validate our findings. Although WGS data was only available on a subset of our participants, it: a) confirmed that our finding of raised urine BMPs in PSP vs. controls was not impacted by the inclusion of PSP cases that are LRRK2 or GBA carriers, although it should be noted that dedicated Gauchian genotyping (**23**) was not available for PSP cases; b) validated the LGC genotyping results that were used to do the pQTL analyses.

In conclusion, biochemically defined lysosomal dysfunction is evident in PSP. We have also shown a pQTL effect of rs76904798 on CSF LRRK2 levels as well as baseline monocyte total LRRK2 levels predicting 1 year change in PSP rating scale scores. Altogether, these findings suggest that genetic and biochemical stratification may identify PSP patients that would benefit from LRRK2-targetting therapies.

## Acknowledgements

We thank the patients and their families for study participation. The PROSPECT-UK study is supported by the PSP Association, CBD Solutions, and the Medical Research Council.

## Author roles

Design: EMS, EJ.

Execution: All authors.

Data analysis: LKN, JF, EMS, EJ

Writing of first draft: LKN, EJ

Editing of final version of the manuscript: All authors.

## Financial disclosures

DPV is supported by CBD Solutions. RR is supported by Aligning Science Across Parkinson’s. AQ is supported by Aligning Science Across Parkinson’s. BCPG is supported in part by an NIHR grant (NIHR207984): the views expressed are those of the authors and not necessarily those of the National Institute for Health and Care Research [NIHR] or the Department of Health and Social Care. CK is supported by the Multiple System Atrophy Trust and Parkinson’s UK. JBR is supported by the Medical Research Council (MC_UU_00030/14; MR/T033371/1), the PSP Association, Cambridge Centre for Parkinson-plus, the NIHR Clinical Research Network, and the NIHR Cambridge Biomedical Research Centre (NIHR203312: the views expressed are those of the authors and not necessarily those of the National Institute for Health and Care Research [NIHR] or the Department of Health and Social Care). HRM is supported by Parkinson’s UK, Cure Parkinson’s Trust, PSP Association, NIHR-MRC (ExPRESS study grant), and The Michael J. Fox Foundation. EJ is supported by a Medical Research Council Clinician Scientist Fellowship (UKRI1388) and grants from the PSP Association (PSPA2023/PROJECTGRANT001) and CurePSP (681-2022/06).

## Declaration of interests

BCPG’s salary is paid by University Hospital Southampton. In the past 12 months BCPG has received honoraria from the neurology masterclass and GEC, grants from the PSP Association, and paid consultancy from ImmunoBrain. BCPG serves as a trustee and is on the research committee for the PSP Association. CK is employed by Northern Care Alliance NHS Foundation Trust. In the past 12 months CK has received speaker honoraria from Neurology Academy, Ipsen, and Britannia Pharmaceuticals. MTMH receives payment for Advisory Board attendance/consultancy from Helicon, NeuHealth Digital, Roche and Manus Neurodynamica, and is an academic founder and shareholder of NeuHealth Digital Ltd. JBR is employed by Cambridge University with academic grants from AstraZeneca, Lilly, GSK, and Janssen, and paid consultancy for Asceneuron, Astex, Astronautx, Alector, Booster Therapeutics, Clinical Ink, Curasen, CumulusNeuro, Eisai, Ferrer, ICG, Invicro, Prevail, in the past 12 months unrelated to the current work. HRM is employed by UCL. In the past 12 months HRM reports paid consultancy from Roche, Aprinoia, AI Therapeutics and Amylyx; lecture fees or honoraria from BMJ, Kyowa Kirin, and the Movement Disorders Society. HRM is a co-applicant on a patent application related to C9ORF72 (Method for diagnosing a neurodegenerative disease; PCT/GB2012/052140). In the past 12 months EJ reports paid consultancy from Novartis.

All authors report no financial disclosures or conflicts of interest regarding this study.

This work was funded by CurePSP, PSP Association, the Medical Research Council, and the National Institute for Health and Care Research.

## Notes

### Competing Interest Statement

The authors have declared no competing interest.

### Author Declarations

The Queen Square Research Ethics Committee gave ethical approval for this work (14/LO/1575).

## References

(1) Dickson DW. Neuropathology of Parkinson’s disease and parkinsonism. Cold Spring Harb Perspect Med. 2025: a041610 (online ahead of print).

(2) Sosero YL, Gan-Or Z. LRRK2 and Parkinson’s disease: from genetics to targeted therapy. Ann Clin Transl Neurol. 2023; 10(6): 850–864.

(3) Nalls MA, Blauwendraat C, Vallerga CL, et al. Identification of novel risk loci, causal insights, and heritable risk for Parkinson’s disease: a meta-analysis of genome-wide association studies. Lancet Neurol. 2019; 18(12): 1091–1102.

(4) Sanchez-Contreras M, Heckman MG, Tacik P, et al. Study of LRRK2 variation in tauopathy: progressive supranuclear palsy and corticobasal degeneration. Mov Disord. 2017; 32(1): 115–123.

(5) Jabbari E, Koga S, Valentino RR, et al. Genetic determinants of survival in progressive supranuclear palsy: a genome-wide association study. Lancet Neurology 2021; 20(2): 107–116.

(6) Henderson MX, Sengupta M, Trojanowski JQ, et al. Alzheimer’s disease tau is a prominent pathology in LRRK2 Parkinson’s disease. Acta Neuropathol Commun. 2019; 7(1): 183.

(7) Siderowf A, Concha-Marambio L, Lafontant DE, et al. Assessment of heterogeneity among participants in the Parkinson’s Progression Markers Initiative cohort using α-synuclein seed amplification: a cross-sectional study. Lancet Neurology 2023; 22(5): 407–417.

(8) Alessi DR, Sammler E. LRRK2 kinase in Parkinson’s disease. Science 2018; 360(6384): 36–37.

(9) Taymans J-M, Fell M, Greenamyre T, et al. Perspective on the current state of the LRRK2 field. NPJ Parkinsons Dis. 2023; 9(1): 104.

(10) Fan Y, Howden AJM, Sarhan AR, et al. Interrogating Parkinson’s disease LRRK2 kinase pathway activity by assessing Rab10 phosphorylation in human neutrophils. Biochem J. 2018; 475(1): 23–44.

(11) Fan Y, Nirujogi RS, Garrido A, et al. R1441G but not G2019S mutation enhances LRRK2 mediated Rab10 phosphorylation in human peripheral blood neutrophils. Acta Neuropathol. 2021; 142(3): 475–494.

(12) Alcalay RN, Hsieh F, Tengstrand E, et al. Higher urine bis(monoacylglycerol) phosphate levels in LRRK2 G2019S mutation carriers: implications for therapeutic development. Mov Disord. 2020; 35(1): 134–141.

(13) Merchant KM, Simuni T, Fedler J, et al. LRRK2 and GBA1 variant carriers have higher urinary bis(monoacylglycerol) phosphate concentrations in PPMI cohorts. NPJ Parkinsons Dis. 2023; 9(1): 30.

(14) Hoglinger GU, Respondek G, Stamelou M, et al. Clinical diagnosis of progressive supranuclear palsy: The movement disorder society. Mov Disord. 2017; 32(6): 853–864.

(15) Jabbari E, Holland N, Chelban V, et al. Diagnosis across the spectrum of progressive supranuclear palsy and corticobasal syndrome. JAMA Neurol 2020; 77(3): 377–387.

(16) Bandres-Ciga S, Faghri F, Majounie E, et al. NeuroBooster Array: a genome-wide genotyping platform to study neurological disorders across diverse populations. Mov Disord. 2024; 39(11): 2039–2048.

(17) Hochberg Y, Benjamini Y. More powerful procedures for multiple significance testing. Stat Med. 1990; 9(7): 811–818.

(18) Buck SA, Malankhanova T, Strader S, et al. LRRK2 kinase-mediated accumulation of lysosome-associated phospho-Rabs in tauopathies and synucleinopathies. Acta Neuropathol. 2025; 150(1): 44.

(19) Cao W, Zheng H. Peripheral immune system in aging and Alzheimer’s disease. Mol Neurodegener. 2018; 13(1): 51.

(20) Evans LD, Strano A, Tuck E, et al. Tau uptake by human neurons depends on receptor LRP1 and kinase LRRK2. EMBO J. 2025: online ahead of print.

(21) Langston RG, Beilina A, Reed X, et al. Association of a common genetic variant with Parkinson’s disease is mediated by microglia. Sci Transl Med. 2022; 14(655): eabp8869.

(22) Meikle PJ, Duplock S, Blacklock D, et al. Effect of lysosomal storage on bis(monoacylglycero) phosphate. Biochem J. 2008; 411(1): 71–78.

(23) Toffoli M, Chen X, Sedlazeck FJ, et al. Comprehensive short and long read sequencing analysis for the Gaucher and Parkinson’s disease-associated GBA gene. Commun Biol. 2022; 5(1): 670.

