## Supplementary material for "Biomarkers of LRRK2 and lysosomal dysfunction in Progressive Supranuclear Palsy"

| **Section** | | **Page** |
| --- | --- | --- |
| 1) | Methods: Immunoblotting | 2 |
| 2) | Table 1: PD/Parkinsonism genes screened for pathogenic variants using WGS and NeuroBooster Array data | 3 |
| 3) | Figure 1: PSP vs. control quantitative immunoblotting plots for DMSO and MLi-2 treated neutrophil and monocyte samples | 4 |
| 4) | Table 2: PSP vs. control comparisons of urine total di-22:6 and di-18:1 BMP levels and associated isoforms | 5 |
| 5) | Table 3: Blood (neutrophil and monocyte) and CSF total LRRK2 and pRab10 levels in PSP and control groups stratified by rs2242367 and rs76904798 genotype status | 6 |
| 6)  7) | Figure 2: Heatmap of Spearman’s rho biomarker correlations in the control group  Table 4: Linear regression models using baseline levels of neutrophil, monocyte, urine and CSF measures to predict 1 year change in the PSP rating scale score | 7  8 |

**Methods: Immunoblotting**

In preparation for multiplexed immunoblotting, the protein concentrations of all cleared neutrophil and monocyte lysates were determined via the BCA assay, and samples were made up to a protein concentration of 2 mg/ml in 4x NuPAGE LDS sample buffer supplemented with 5% (v/v) β-mercaptoethanol. For each sample, 20 μg of protein was loaded in duplicates onto 20-well commercial NuPAGE 4-12% Bis-Tris gels and electrophoresed in MOPS SDS running buffer at 90V for 15 minutes and then 140V until the dye-front was at the bottom of the gel. The electrophoresed proteins were transferred from the gels onto nitrocellulose membranes using wet transfer at 90V for 90 minutes in transfer buffer (48 mM Tris, 39 mM glycine, 20% (v/v) methanol). Following transfer, the membranes were stained with Ponceau S and cut to separate the proteins of interest (LRRK2, Rab10, and the housekeeping protein, GAPDH). The ponceau strain was washed off with TBS-T (20 mM Tris, 150 mM NaCl, 0.2% (v/v) Tween20), after which the membranes were blocked at room-temperature for 1 hour in 5% (w/v) milk in TBS-T. Following washing in TBS-T, the membranes were incubated overnight at 4 °C in primary antibodies (multiplexed total LRRK2 and pSer935 LRRK2, multiplexed total Rab10 and pThr73 Rab10, and GAPDH) diluted in a TBS-T buffer with 5% (w/v) BSA and 0.02% sodium azide. From this point forward, the membranes were kept in the dark. The next day, the membranes were washed 3x5 minutes in TBS-T and incubated in secondary antibodies diluted in TBS-T (LI-COR IRDye 800CW and/or IRDye 680RD) at room temperature for 1 hour. The membranes were washed 3x10 minutes in TBS-T and imaged on a LI-COR Odyssey CLx scanner. Finally, the signal intensities were quantified in ImageStudio.

**Table 1**

| ATP13A2 | LYST | SLC30A10 |
| --- | --- | --- |
| ATP1A3 | MAPT | SLC39A14 |
| C19orf12 | OPA3 | SLC6A3 |
| CSF1R | PANK2 | SNCA |
| DCTN1 | PARK7 | SPG11 |
| DNAJC6 | PDGFB | SPR |
| FBXO7 | PINK1 | SYNJ1 |
| FTL | PLA2G6 | TH |
| GBA | PRKN | TUBB4A |
| GCH1 | PRKRA | VPS13A |
| GRN | PTRHD1 | VPS35 |
| LRRK2 | RAB39B | WDR45 |

**Table 1:** PD/Parkinsonism genes screened for pathogenic variants using WGS and NeuroBooster Array data.

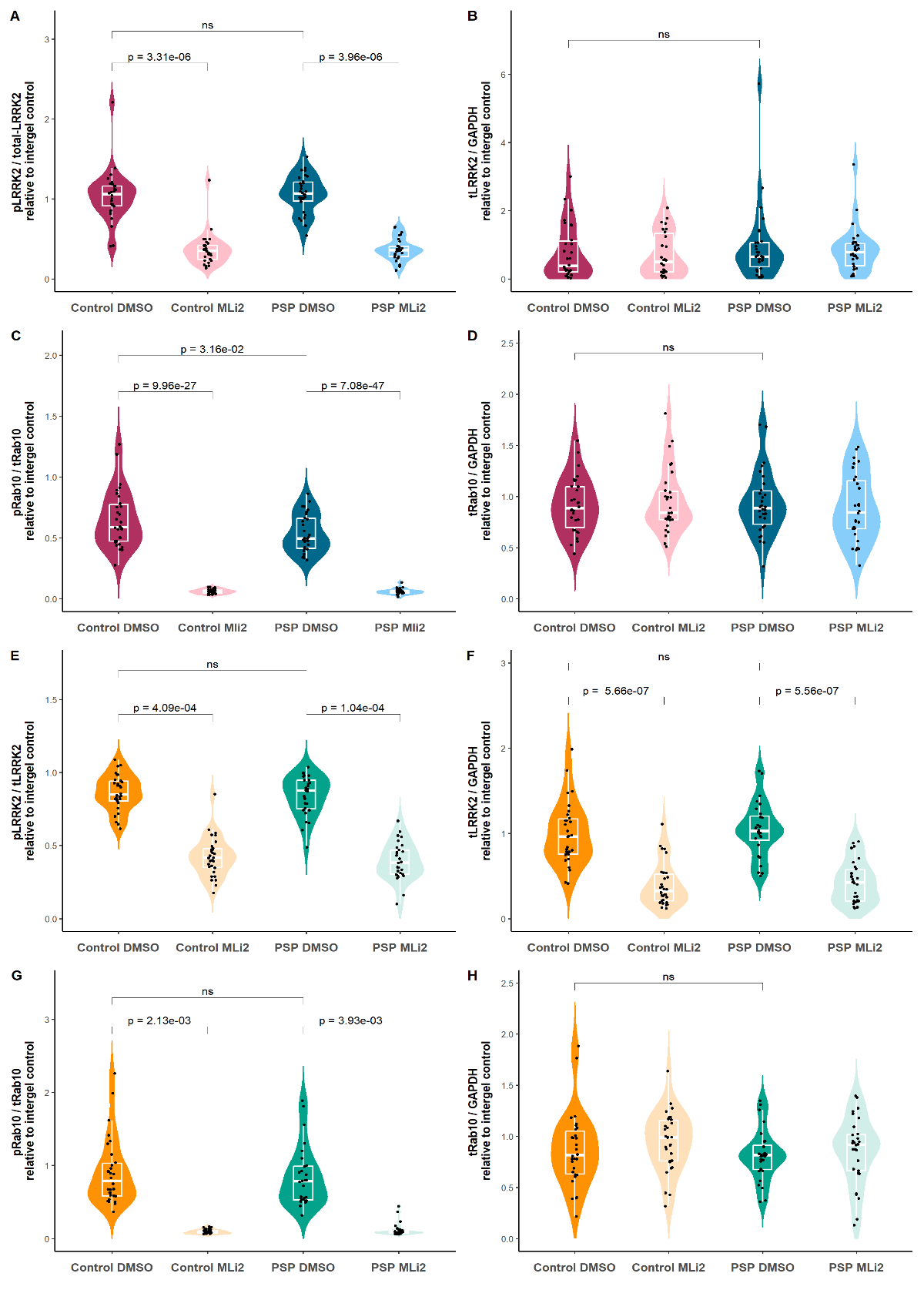
**Figure 1**

**Figure 1:** PSP vs. control quantitative immunoblotting plots for DMSO and MLi-2 treated samples regarding: a) neutrophil phosphorylated LRRK2 levels normalised by total LRRK2; b) neutrophil total LRRK2 levels normalised by GAPDH; c) neutrophil phosphorylated Rab10 levels normalised by total Rab10; d) neutrophil total Rab10 levels normalised by GAPDH; e) monocyte phosphorylated LRRK2 levels normalised by total LRRK2; f) monocyte total LRRK2 levels normalised by GAPDH; g) monocyte phosphorylated Rab10 levels normalised by total Rab10; h) monocyte total Rab10 levels normalised by GAPDH. Group comparisons done using logistic regression that adjusted for sex and age at testing.

**Table 2**

| **Logistic regression results** | | | |
| --- | --- | --- | --- |
|  | OR | 95% CI | p-value |
| Total di-18:1-BMP | 1.06 | (0.94, 1.19) | 0.33 |
| di.18.1.BMP.2.2 | 1.02 | (0.89, 1.18) | 0.74 |
| di.18.1.BMP.2.3 | 1.79 | (0.86, 3.70) | 0.12 |
| di.18.1.BMP.3.3 | 16.30 | (2.01, 131.00) | **0.01** |
| Total.di.22.6.BMP | 1.04 | (1.00, 1.08) | **0.04** |
| di.22.6.BMP.2.2 | 1.04 | (0.99, 1.10) | 0.10 |
| di.22.6.BMP.2.3 | 1.14 | (1.00, 1.30) | 0.05 |
| di.22.6.BMP.3.3 | 1.35 | (0.98, 1.84) | 0.06 |

**Table 2:** PSP vs. control comparisons of urine total di-22:6 and di-18:1 BMP levels and their associated isoforms. Group comparisons done using logistic regression that adjusted for sex and age at testing.

**Table 3**

| **SNP: rs2242367** | | | | | |
| --- | --- | --- | --- | --- | --- |
|  | Group | OR | 95% CI | | p-value |
| CSF LRRK2 | PSP | 1.12 | (0.99, 1.26) | | 0.08 |
| CSF pRab10 | PSP | 0.52 | (0.09, 2.91) | | 0.45 |
| Neutrophil tLRRK2/GAPDH | PSP | 1.79 | (0.67, 4.82) | | 0.25 |
|  | Control | 8.09 | (0.99, 65.70) | | 0.05 |
| Neutrophil  pRab10/tRab10 | PSP  Control | 9.81  4.29 | (0.01, 9476.00)  (0.04, 520.00) | | 0.52  0.55 |
| Monocyte tLRRK2/GAPDH | PSP | 0.40 | (0.01, 11.90) | | 0.59 |
|  | Control | 0.22 | (0.02, 2.41) | | 0.22 |
| Monocyte  pRab10/tRab10 | PSP  Control | 1.02  2.09 | (0.03, 41.40)  (0.27, 16.50) | | 0.99  0.48 |
| **SNP: rs76904798** | | | | | |
|  | Group | OR | | 95% CI | p-value |
| CSF LRRK2 | PSP | 1.15 | | (1.02, 1.29) | **0.02** |
| CSF pRab10 | PSP | 1.66 | | (0.31, 8.83) | 0.55 |
| Neutrophil tLRRK2/GAPDH | PSP | 0.78 | | (0.32, 1.93) | 0.59 |
|  | Control | 0.21 | | (0.01, 5.94) | 0.36 |
| Neutrophil  pRab10/tRab10 | PSP  Control | 0.04  0.03 | | (0.00, 21.30)  (0.00, 52.20) | 0.31  0.36 |
| Monocyte tLRRK2/GAPDH | PSP | 0.64 | | (0.02, 19.00) | 0.80 |
|  | Control | 1.53 | | (0.08, 30.30) | 0.78 |
| Monocyte  pRab10/tRab10 | PSP  Control | 0.73  0.12 | | (0.02, 25.30)  (0.02, 8.45) | 0.86  0.32 |

**Table 3:** Blood (neutrophil and monocyte) and CSF total LRRK2 and pRab10 levels in PSP and control groups stratified by rs2242367 and rs76904798 genotype status. Genotype group comparisons done using logistic regression that adjusted for sex, age and disease duration at testing (control group comparisons adjusted for sex and age at testing) with GG (rs2242367) and CC (rs76904798) as reference groups. OR = odds ratio; 2.5%/97.5% = confidence interval of the odds ratio. CSF LRRK2 levels in the control group were not included due to the low number of observations.

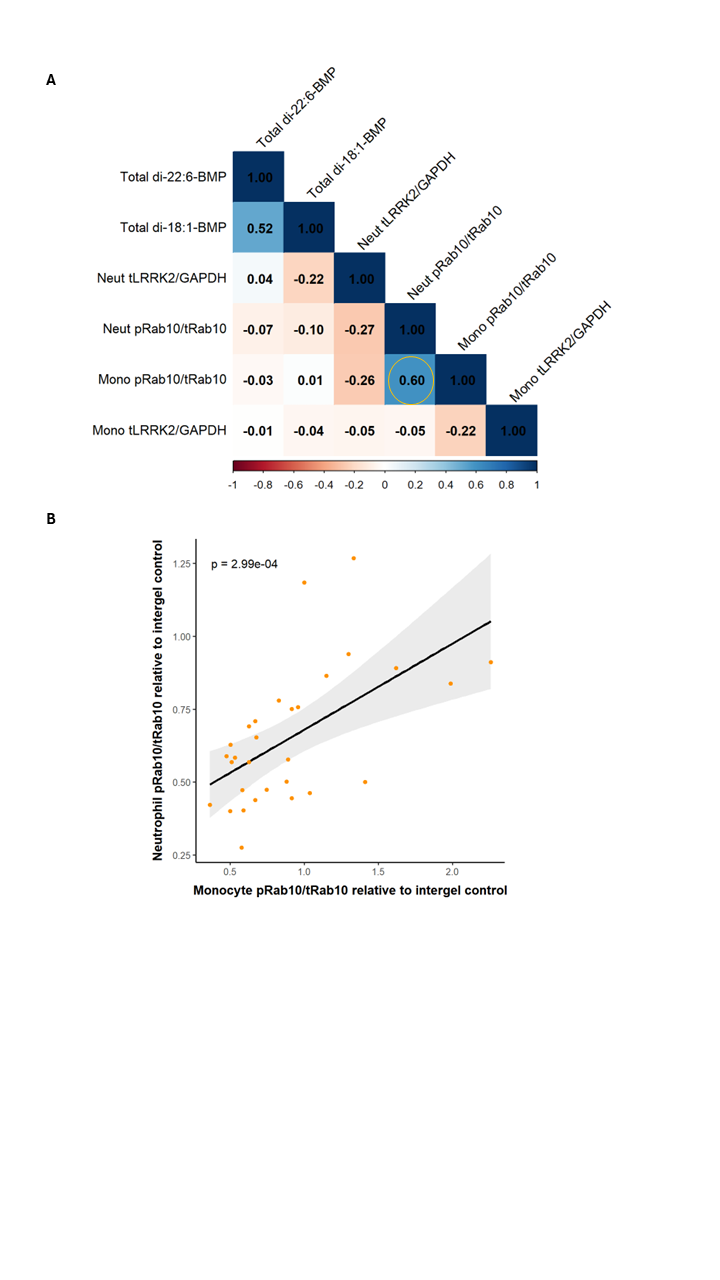
**Figure 2**

**Figure 2:** a) Heatmap of Spearman’s rho biomarker correlations in the control group with r values highlighted. Circle represents correlation that was also significant in the linear regression analysis; b) neutrophil pRab10 vs. monocyte pRab10 linear plot. A linear regression analysis that adjusted for sex and age at testing was used to generate the p-value.

**Table 4**

|  | Variable | Estimate | 95% CI | p-value |
| --- | --- | --- | --- | --- |
| PSPRS change  (1 year) | Neutrophil tLRRK2/GAPDH | -6.61 | (-11.40, -1.78) | 0.03 |
|  | Neutrophil pRab10/tRab10 | 3.13 | (-25.90, 32.10) | 0.84 |
|  | Monocyte tLRRK2/GAPDH | 17.80 | (8.38, 27.30) | **0.008** |
|  | Monocyte pRab10/tRab10 | 0.59 | (-11.90, 13.10) | 0.93 |
|  | CSF LRRK2 | 0.54 | (-0.25, 1.33) | 0.19 |
|  | CSF pRab10 | 8.51 | (-3.07, 20.10) | 0.16 |
|  | Total di-18:1-BMP | -0.19 | (-0.81, 0.44) | 0.58 |
|  | Total di-22:6-BMP | 0.12 | (-0.21, 0.45) | 0.50 |

**Table 4:** Linear regression models using baseline levels of neutrophil, monocyte, urine and CSF measures to predict 1 year change in clinical rating scale scores in PSP adjusted for sex, age and disease duration at baseline testing. Confidence interval = CI, PSP rating scale = PSPRS, Movement Disorder Society-Unified Parkinson’s Disease Rating Scale part III = MDS-UPDRS III, Montreal Cognitive Assessment = MoCA. Benjamini-Hochberg corrected p-value significance threshold < 0.009.
